# Computational Image Analysis Techniques, Programming Languages and Software Platforms Used in Cancer Research: A Scoping Review

**DOI:** 10.1101/2022.04.26.22274298

**Authors:** Youssef Arafat, Constantino Carlos Reyes-Aldasoro

## Abstract

**Background:** Cancer-related research, as indicated by the number of entries in Medline, the National Library of Medicine of the USA, has dominated the medical literature. An important component of this research is based on the use of computational techniques to analyse the data produced by the many acquisition modalities. This paper presents a review of the computational image analysis techniques that have been applied to cancer. The review was performed through automated mining of Medline/PubMed entries with a combination of keywords. In addition, the programming languages and software platforms through which these techniques are applied were also reviewed.

**Methods:** Automatic mining of Medline/PubMed was performed with a series of specific keywords that identified different computational techniques. These keywords focused on traditional image processing and computer vision techniques, machine learning techniques, deep learning techniques, programming languages and software platforms.

**Results:** The entries related to traditional image processing and computer vision techniques have decreased at the same time that machine learning and deep learning have increased significantly. Within deep learning, the keyword that returned the highest number of entries was *convolutional neural network*. Within the programming languages and software environments, Fiji and ImageJ were the most popular, followed by Matlab, R, and Python. Within the more specialised softwares, QuPath has had a sharp growth overtaking other platforms like ICY and CellProfiler.

**Conclusions:** The techniques of artificial intelligence techniques and deep learning have grown to overtake most other image analysis techniques and the trend at which they grow is still rising. The most used technique has been convolutional neural networks, commonly used to analyse and classify images. All the code related to this work is available through GitHub: https://github.com/youssefarafat/Scoping-Review.

## 1 Introduction

Cancer has dominated the medical literature. At the time of writing of this paper (June 2022), there were 4,633,885 Cancer-related entries from the total 34,216,925, which represented 13% of all entries in PubMed (https://www.ncbi.nlm.nih.gov/pubmed), the search engine of the United States National Library of Medicine MEDLINE. These entries have risen from 6% of all yearly entries recorded in PubMed in the 1950s, to more than 16% seventy years later [19]. Among many technological innovations that have had an impact in medicine in general, and in Cancer in particular, computational data analysis techniques have been important, especially as new imaging technologies provide more and more data every year. Diagnostic images acquired with Magnetic Resonance Imaging [1], Computed Tomography [27], Ultrasound [8], and pathology staining [12, 25] are routinely obtained and analysed, traditionally by experts that trained for many years to provide accurate diagnosis. In addition, computational techniques have been used to support decision-making [28], preoperative planning [23] and predict survival based on the analysis of the images acquired [7].

In recent years, the computational techniques included under the umbrella term of Artificial Intelligence have grown at a very fast rate, but before that, a rich body of techniques developed under the areas of Image Processing and Computer Vision were applied to analyse data from a variety of Cancer data sets. Techniques like Fourier analysis [4], mathematical morphology [22], and fractals [13] among many others have been applied with success in the analysis of Cancer treatments or drug development [15, 16, 18]. The question that arises is then, are the artificial intelligence techniques displacing all the previous image processing and computer vision techniques as applied to the analysis of Cancer images?

In this paper, a review of the data analysis techniques that have been applied to Cancer data analysis is presented. Since the objective of the review is to map the literature, assess the volume of work, as reflected by the number of entries in PubMed, rather than compare outcomes or perform a precise systematic review and synthesis of the evidence [17, 26], a scoping review, rather than a systematic review was performed. The review utilised data mining techniques to extract the entries related to different techniques that have been applied to the analysis of Cancer images. A series of keywords were used in combination to select different techniques, and their yearly entries in PubMed. In addition, the tools through which these techniques were applied, i.e. the software platforms or programming languages were also reviewed.

## 2 Materials and Methods

The scoping review followed the Preferred Reporting Items for Systematic Reviews and Meta-Analyses (PRISMA) [11, 14] and the extension for Scoping Reviews (PRISMAScR) guidelines [17, 26] and included the following stages: (1) Identify the research questions, (2) Identify data source and the combination of keywords that would retrieve the relevant entries, (3) Mine the number of entries per year of publication, (4) Display the trends graphically, (5) Interpret the findings. One notable difference of this review is that the screening and eligibility steps were performed automatically by sequentially adding a series of specific keywords to a basic search instead of excluding entries manually or with criteria other than the presence of keywords. The inclusion was solely based on the specific keywords being present in the query as described below.

### 2.1 Research Questions

The following questions were considered: *Which computational techniques of those considered traditional (i*.*e. not deep learning) and deep learning have been most widely employed in the analysis of images related to Cancer data sets?. Which tools have been most widely used to apply the computational techniques previously mentioned?*

### 2.2 Keywords and Data Source

The only database considered in this review was Medline/PubMed, which has been considered to be “the most widely used database of biomedical literature” [24]. The first keyword selected in the queries was *Cancer*, which returned more than 4 million entries (June 2022). With the addition of *Pathology* and a logical AND, entries were reduced to 1,668,363. To focus on image processing/analysis, *images AND imaging* were added and these reduced the entries to 283,702. Of these, 261,058 were published between 1990 and 2022, which were the basic sample to which particular keywords were further added.

Three groups of specific keywords were used to explore the landscape: traditional Image Processing and Computer Vision, Machine Learning and Deep Learning Techniques. The specific keywords are listed in Table 1.

**Table 1.** Specific keywords used in the search strategy to explore Image Processing and Computer Vision, the Machine Learning and Deep Learning Techniques.

| Image Processing /<br>Computer Vision | Machine Learning | Deep Learning |
| --- | --- | --- |
| texture analysis | Classification | Deep belief |
| Fourier | Regression | Recurrent neural networks |
| geometric | Dimensionality reduction | Auto-Encoders |
| tracing | Ensemble learning | Multilayer perceptron |
| linear discriminant analysis | Reinforcement learning | Generative adversarial network |
| thresholding | Supervised learning | Convolutional neural network |
| feature extraction | Bayesian | U-Net |
| tracking | Decision tree | Transformer |
| clustering | Linear classifier | Fully convolutional |
| scale space | Unsupervised learning | ResNet (Residual |
| hessian | Artificial neural networks | neural network ) |
| self-organizing | Hierarchical clustering | VGG |
| region growing | Cluster analysis | Mask-RCNN (Region-Based |
| mutual information | Anomaly detection | Convolutional Neural Networks ) |
| wavelet | Semi-supervised learning | LSTM (Long short-term |
| multiresolution | Deep learning | memory) |
| principal component analysis | Support Vector Machines | GoogleNet |
| filtering | Naive Bayes | AlexNet |
| active contour | Nearest Neighbor | Inception-v3 |
| fractal | Discriminant Analysis | DenseNet |
| linear regression | K-Means | Inception-ResNet-v2. |
| ensemble | Hidden Markov Model |  |
| transfer learning | Feature Selection |  |
| convolutional neural | Feature Engineering |  |
| machine learning | Random Forest. |  |
| deep learning |  |  |

The first group of specific keywords were used to investigate the traditional image processing / computer vision. Some of these keywords were specific enough to be used as a single word, like *Fractal*. Others required a combination of 2 or more terms, for instance, texture could be related to the texture of a sample and not to the specific technique of analysis and thus *texture analysis* was used.

Ambiguity is still possible, for instance, texture can be explored with Fourier transforms and an author under the name of Fourier could be included in the entries when no Fourier transforms or analysis were applied. However, the results from queries were manually observed and the keywords were refined to minimise these artefacts. The second and third group of keywords were related to Machine Learning and Deep Learning techniques, which have grown significantly in recent years.

The number of entries were analysed per year and in total for the period of analysis. To analyse the entries per year, in addition to absolute number of entries, a relative number was studied. There are 2 ways in which the relative numbers can be obtained. First, as a percentage of the total number of entries of all the keywords. That is, for a given year, the number of entries of all keywords are added and the ratio of the value of each keyword is divided by that total. Second, as a percentage of the total number of entries of the year irrespective of the specific keywords. In this way, if there are entries that do not include any of the keywords will also be counted to the total.

One final set of queries was performed to investigate the programming languages, softwares and environments that have been used for image analysis of cancer and pathological data sets. The most common options within the “point and click” and computational environments (as described in [5]) were considered: the general programming languages *Matlab, R, C* and *Python* and the specialised softwares *QuPath* [2], *CellProfiler* [9], *Fiji* [20], *ImageJ* [21] and *ICY* [3].

Several of these names can be directly keywords, like CellProfiler and QuPath, which referred exclusively to the tools to be investigated and did not imply any ambiguity. However, it was noticed that in some cases, the use of the keyword returned fewer entries than the number of citations to a specific paper. For instance searching for QuPath (https://pubmed.ncbi.nlm.nih.gov/?term=QuPath) returned 86 entries, yet the citations to the original paper describing QuPath (https://pubmed.ncbi.nlm.nih.gov/?linkname=pubmed_pubmed_citedin&from_uid=29203879) returned 780 entries. In other cases, like Matlab or R, would return entries as a keyword, but there was no original paper or reference that could be used.

Thus, two different search strategies were performed: the entries related to *Matlab, R, C and Python* were extracted directly through searches with specialised keywords in the URL. The entries related to *Fiji, ImageJ, ICY, QuPath* and *CellProfiler* were extracted from the citations. Whilst these comparisons are not exactly *like with like*, they provide a panoramic view of the tools used in the research reported in PubMed. In both cases the entries per year were available through the field *yearCounts* as described below in Mining Strategy.

To define the specific keywords of the programming languages the following considerations were taken into account. *Matlab* could refer to the programming environment of Mathworks and also to a rural area of Bangladesh and thus was queried as *(Matlab)) NOT ((rural) AND (bangladesh))*. The strategy to mine programming languages R and C was more complicated as a single letter cannot be queried effectively in PubMed, thus these keywords were expanded to the following, *((“R project”) OR (“R package”) OR (Rstudio) OR (R/Shiny))* and *(“C programming”) OR (“C language”) OR (“C package”)*. For Python, it was necessary to discard those entries that were related to python snakes and the keywords were formed as follows: *(Python) NOT (snake) NOT (python regius)*.

Whilst *Fiji* was not mined through keywords, this would have a high ambiguity and it would have been necessary to discard the entries related to the country, entries by authors of universities in Fiji, or cities. One search that would reduce these conditions would be: *(Fiji) NOT (suva) NOT (pacific) NOT (Samoa) NOT (Palau) NOT (Nausori)*.

### 2.3 Mining Strategy

To identify the main trends of these analysis techniques, the Medline / PubMed database was queried with a combination of keywords following the methodology previously described [19, 6].

The mining of PubMed was performed with a combination of keywords, which were combined into one Uniform Resource Locator (URL), e.g., a website address in which parameters are used to query the database. The first part of the URL was the address of PubMed (i.e. https://www.ncbi.nlm.nih.gov/pubmed/) and this was followed by search term, which started with *?term=* followed by the concatenation of keywords. Since URLs do not accept certain special characters like quotes or spaces, these need to be converted to the ASCII character set (space = %20, quotes = %22). The first keywords used were: *cancer* and *pathology*. Next, to restrict to those entries related with images, the following was used: *((image) OR (imaging))*. Next, the years of analysis were restricted, initially from 1990 to 2022, and later on, to focus on more recent entries, from 2010 to 2022. The concatenation of keywords with logical AND functions formed the **basic URL** (e.g. https://pubmed.ncbi.nlm.nih.gov/?term=(pathology)%20AND%20(cancer)%20AND%20((image)+OR+(imaging))%20AND%20(2010:2022[dp])) to which then specific keywords (e.g. *Fourier*) added to investigate the research questions.

Year-on-year entries were extracted directly from the query for all keywords as PubMed includes a field called *timelineData* with pairs of values [year, entries] for all the years that have entries. For example, the reader can try to search a term in PubMed, say *Cancer*, display the page source (*view-source*:https://pubmed.ncbi.nlm.nih.gov/?term=cancer) and search within that page for the string *yearCounts*.

All the code used in this paper was developed as Matlab^®^ (The Mathworks™, Natick, USA) functions and Python Jupyter Notebooks, and both are available in the GitHub repository related to this publication (https://github.com/youssefarafat/Scoping-Review). The graphics used in this paper were generated in Matlab.

## 3 Results and Discussion

During the initial period of study, 1990-2022, the analysis techniques that had the higher number of entries were those identified by the keywords: *machine learning, tracking* and *deep learning* with more than 1,500 entries in PubMed. These were followed by five keywords with under 1,000 entries: *linear regression, convolutional neural, Fourier, texture analysis* and *clustering*. On the other side, the keywords which returned the lowest number of entries corresponded to: *mutual information, region growing, multiresolution, self-organizing, hessian* and *scale space* (Fig. 2).

**Fig. 1.**
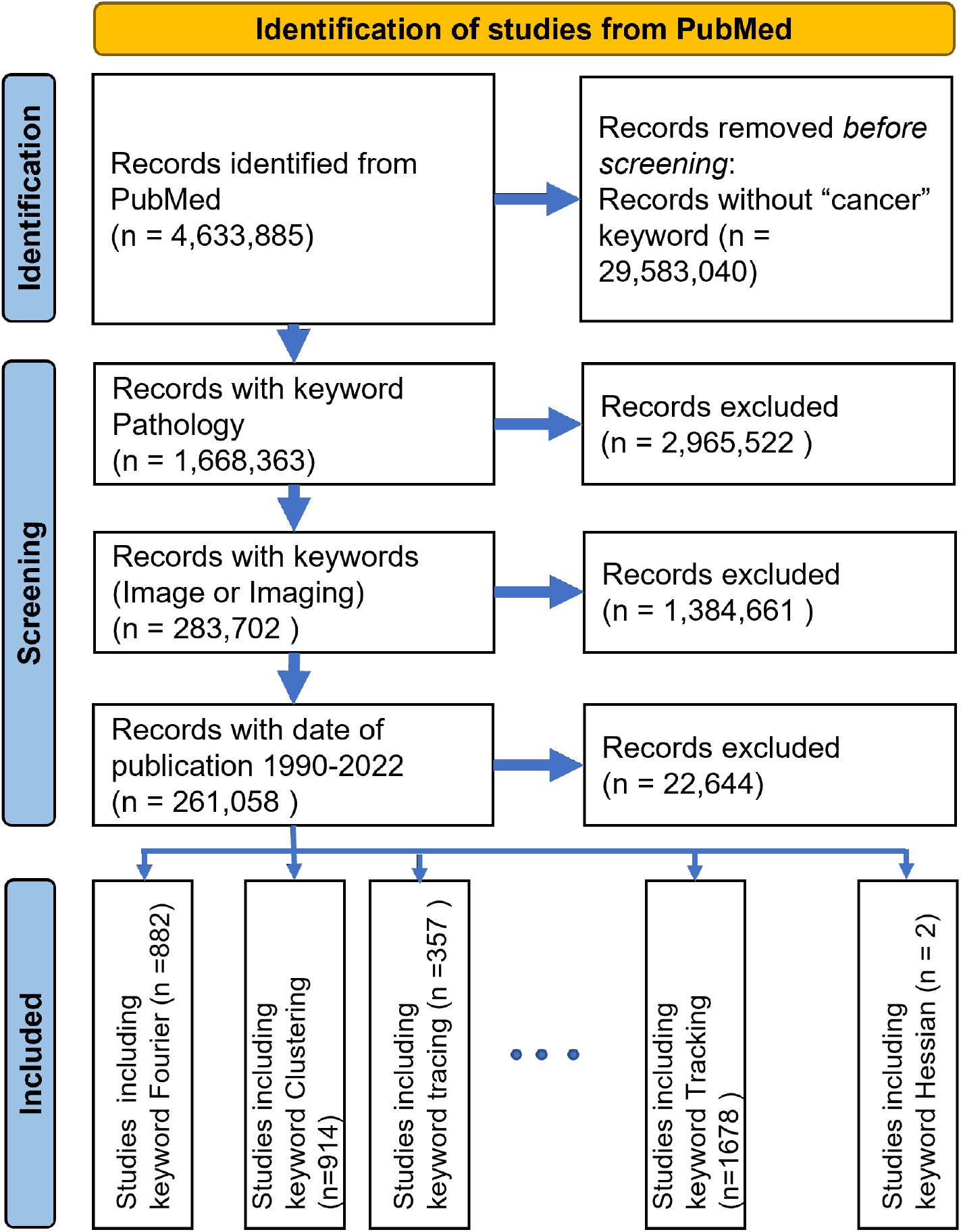
PRISMA statement flowchart illustrating the details of the scoping review. Notably, the inclusion was given by the presence of keywords on PubMed for individual searches. Studies were not read individually.

**Fig. 2.**
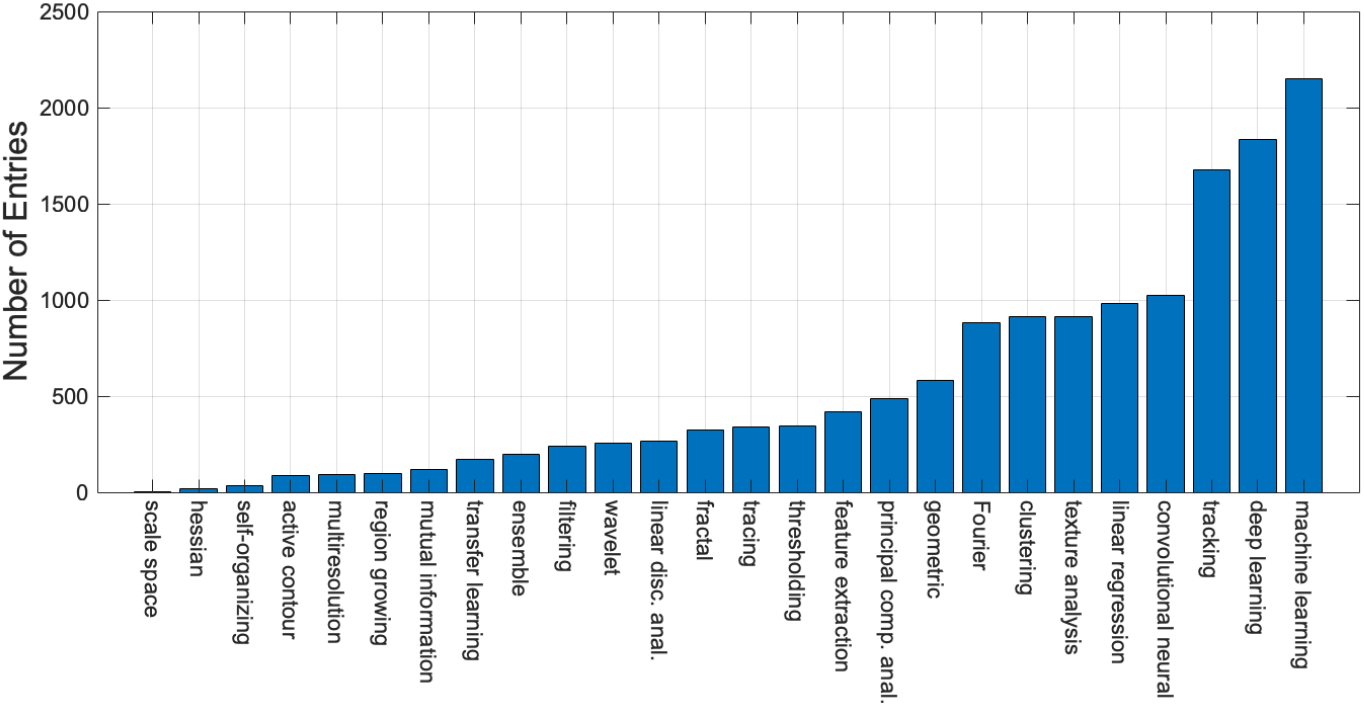
Number of entries in PubMed corresponding to data analysis techniques as determined by specific keywords. The queries were formed with the combination of the following terms (*keyword*) AND (pathology) AND (cancer) AND ((image) OR (imaging)) AND (1990:2022[dp]). This last term corresponds to the date of publication.

The year-on-year analysis of these same keywords showed some interesting trends and a rather different story (Fig. 3). Whilst it would be expected that the number of entries would increase for all keywords, as would be expected since the number of entries in PubMed have grown year on year, from 410,894 entries in 1990 to 1,763,742 entries in 2021 (https://pubmed.ncbi.nlm.nih.gov/?term=(1990[dp]), https://pubmed.ncbi.nlm.nih.gov/?term=(2021[dp])), some cases seem to grow faster. It is clear that some techniques, namely *machine learning, deep learning and convolutional neural*, have accumulated their entries quite recently as opposed to *tracking, linear regression* and *Fourier. Clustering, texture analysis* and *feature extraction* also seem to show an increase, but far less pronounced and difficult to appreciate in this graph. For this reason it is important to observe the trends in relative terms. The ratio of entries of every specific keyword divided by the sum of the entries of all the keywords in every year is shown in Fig. 4. This graph shows the relative contribution of each keyword against the rest for a particular year and emphasises the relative decrease of certain techniques, like *Fourier* and *linear regression*. It should be noticed that ratios of these two techniques were close to 30% in the early 90s. *Tracking* accumulated the majority of its entries in the 2000s but has shown a steep decrease lately.

**Fig. 3.**
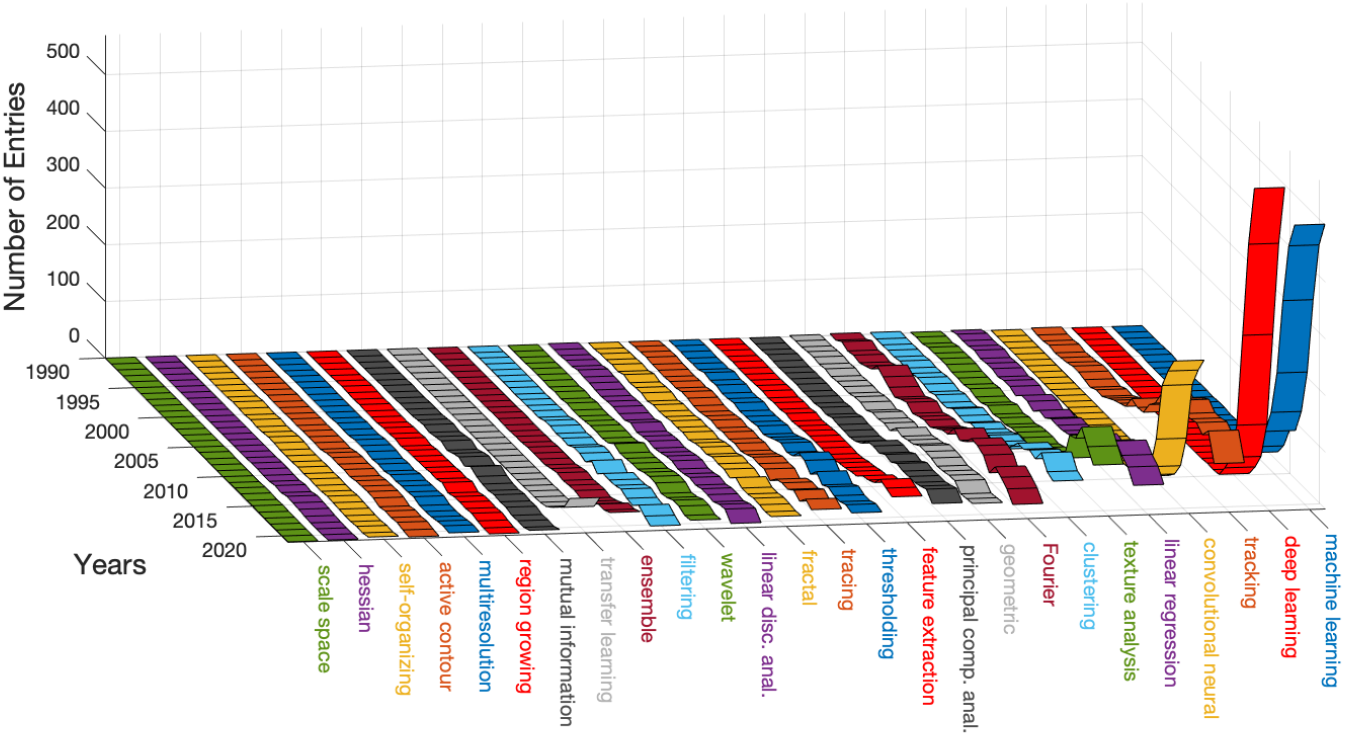
Number of entries in PubMed corresponding to the specific keywords and the date of publication. Colours have been allocated to the ribbons and the names to aid visual discrimination of the computational techniques.

**Fig. 4.**
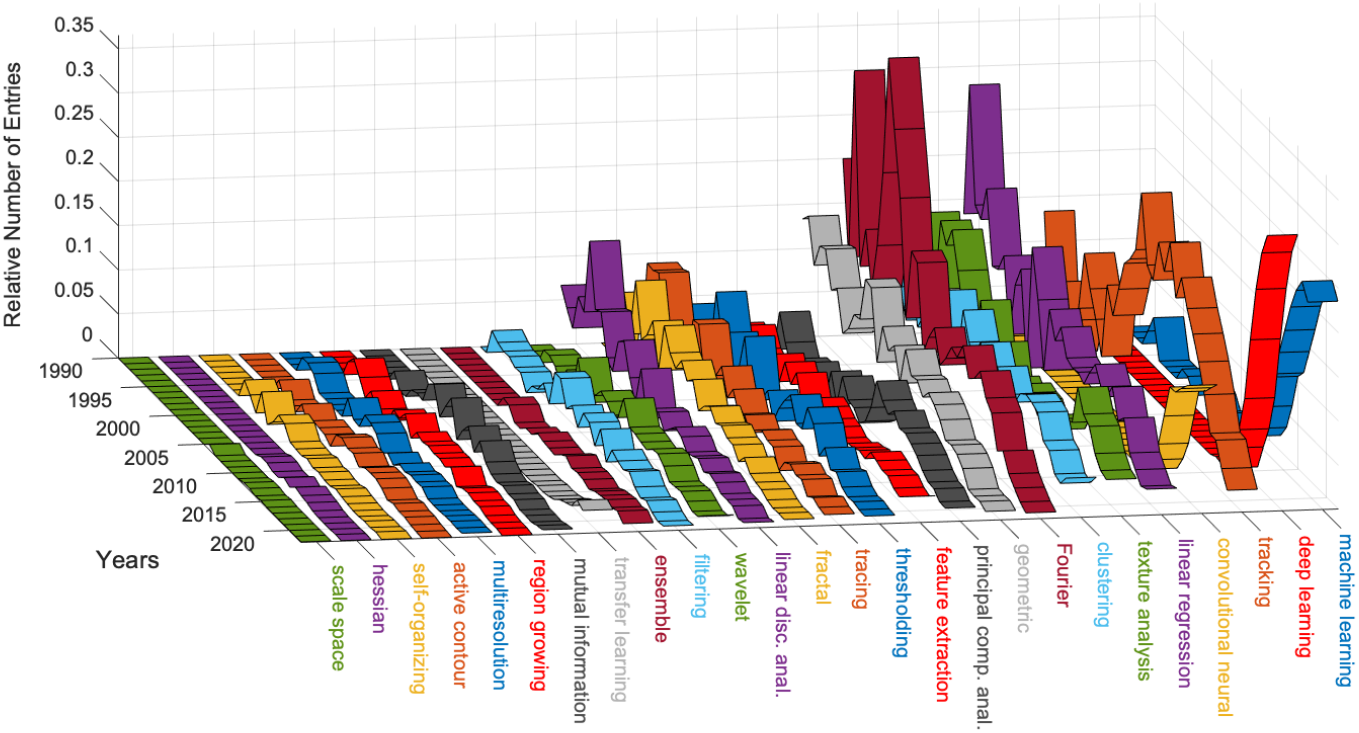
Relative number of entries in PubMed per keyword and date of publication. The number of entries was divided by the sum of all the entries corresponding to the keywords of a given year and thus emphasises relative decrease of certain techniques. Colours have been allocated to the ribbons and the names to aid visual discrimination of the computational technique.

The number of entries for machine learning specific keywords is illustrated in Fig. 5. The first point to notice is that the vertical scale is logarithmic, as the number of entries for *Regression* and *Classification* was much higher than the rest, and this would be expected as these two terms encompass other terms, for instance, classification with random forests. However, these are useful to give perspective to the following entries. *Deep learning* is one order of magnitude above the next keywords; *Feature Selection, Random Forest, Discriminant Analysis* and *Cluster Analysis*. On the lower side, the keywords with fewer entries were *Feature Engineering, Reinforcement learning, Anomaly detection* and *Hidden Markov Model*. It is important to highlight at this stage, that the queries only returned what is indexed in PubMed. If the work of a certain entry did use a technique, say, feature engineering, but this appeared in the methods of the paper, but not in the fields of the PubMed entry, that entry would not be counted in the query. As an example of the data that was mined with the queries, i.e., title, abstract, MESH terms, etc., the following URL https://pubmed.ncbi.nlm.nih.gov/31783023/?format=pubmed corresponds to the work by Lee *et al*. [10].

**Fig. 5.**
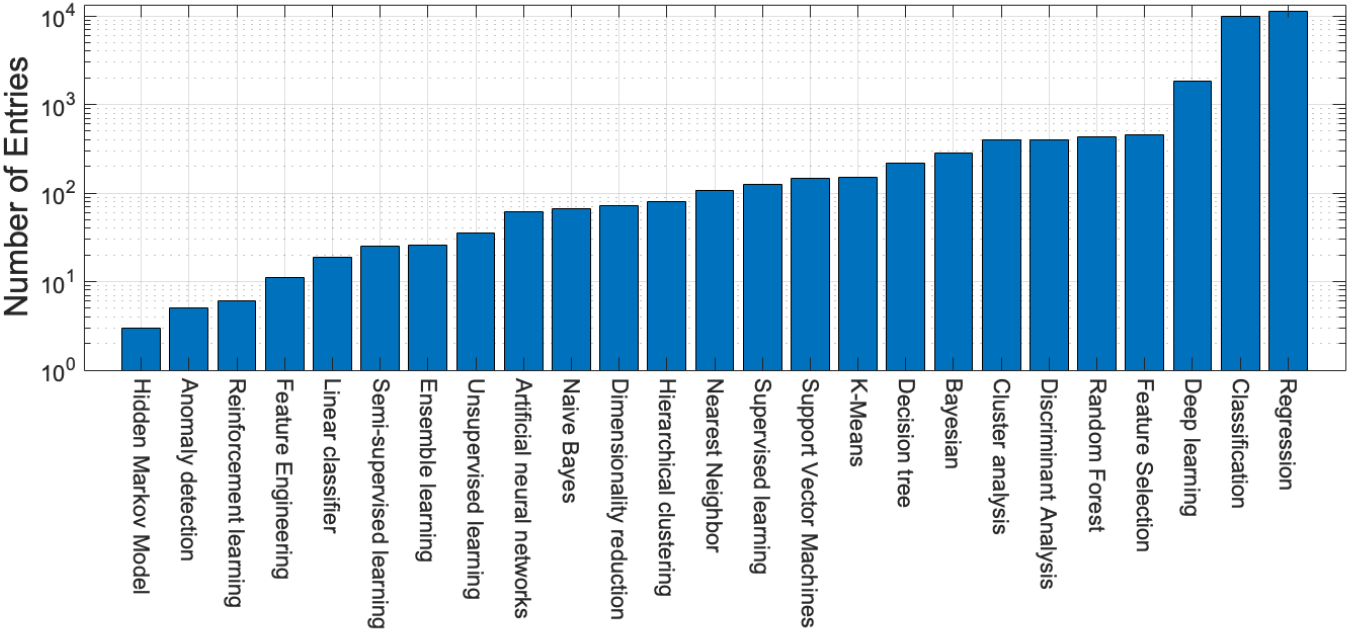
Number of entries in PubMed per specific keyword related to Machine Learning Techniques. The terms “Regression” and “Classification” may include other keywords (e.g., Classification with Random Forest) and thus are more frequent. It can be seen that the term “Deep learning” is one order of magnitude higher than all others. The vertical axis is logarithmic.

When visualised per year, the trends are again very interesting (Fig. 6). In general, the entries corresponding to all specific keywords seem relatively stable, except for one, *deep learning*, which has grown significantly since 2010 and is very close to the entries corresponding to regression and classification. It can be speculated that in a few years, there will be more entries for deep learning than for the other keywords.

**Fig. 6.**
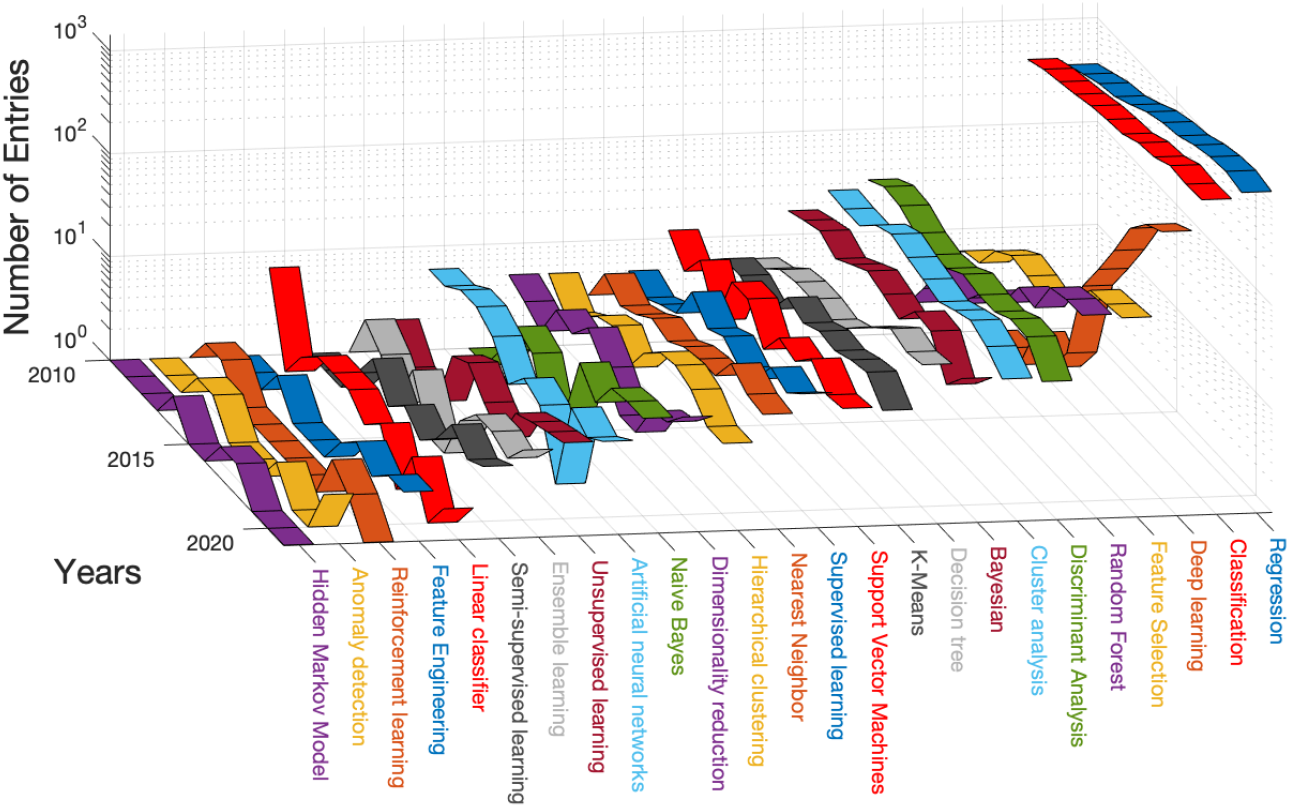
Number of entries in PubMed per year and specific keyword related to Machine Learning Techniques. It should be noticed that entries related most techniques are fairly constant except for “Deep learning” which has increased constantly. It should be noticed that the vertical axis is logarithmic.

The results for specific deep learning keywords are shown in Fig. 7, where the most common term is *convolutional neural network*, followed by *U-Net* and *ResNet*. Convolutional Neural Networks, commonly known as CNNs or Con-vNets, is almost one magnitude higher than all the other methods. The keywords with fewest entries were *Auto-Encoders, Transformers* and *Mask-RCNN*. It should be noticed that some of these terms are rather recent and thus would not be accumulating many entries in PubMed yet.

**Fig. 7.**
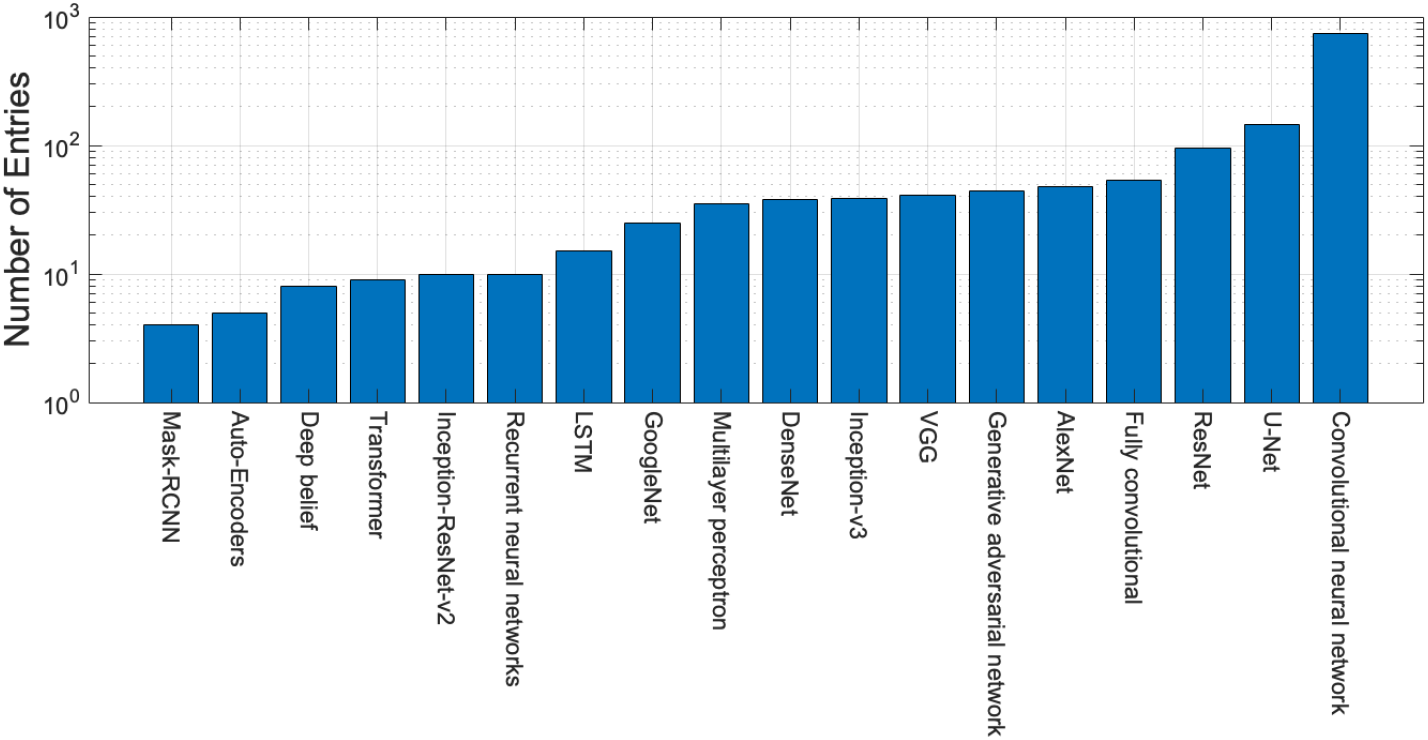
Number of entries in PubMed per specific keyword related to Deep Learning Techniques and some specific architectures like AlexNet, VGG and U-Net. The vertical axis is logarithmic.

The entries corresponding to software and programming languages are shown in Fig. 8. These are shown in two ways, as total number of entries (a) and yearly (b), in both cases the vertical axes are logarithmic. *Fiji* and *ImageJ* returned the highest number of entries followed by *Matlab, R* and *Python*. One order of magnitude below were *QuPath, ICY, CellProfiler*, and this could be expected as the former are general platforms and languages that can be used for a variety of tasks, whilst the latter are more specialised for specific tasks on Quantitative Pathology, Bioimage Informatics and measuring and analysing cell images. As mentioned previously, the queries were performed in different ways and these did not include the keywords cancer, pathology and image analysis, thus they cover all entries indexed in PubMed.

**Fig. 8.**
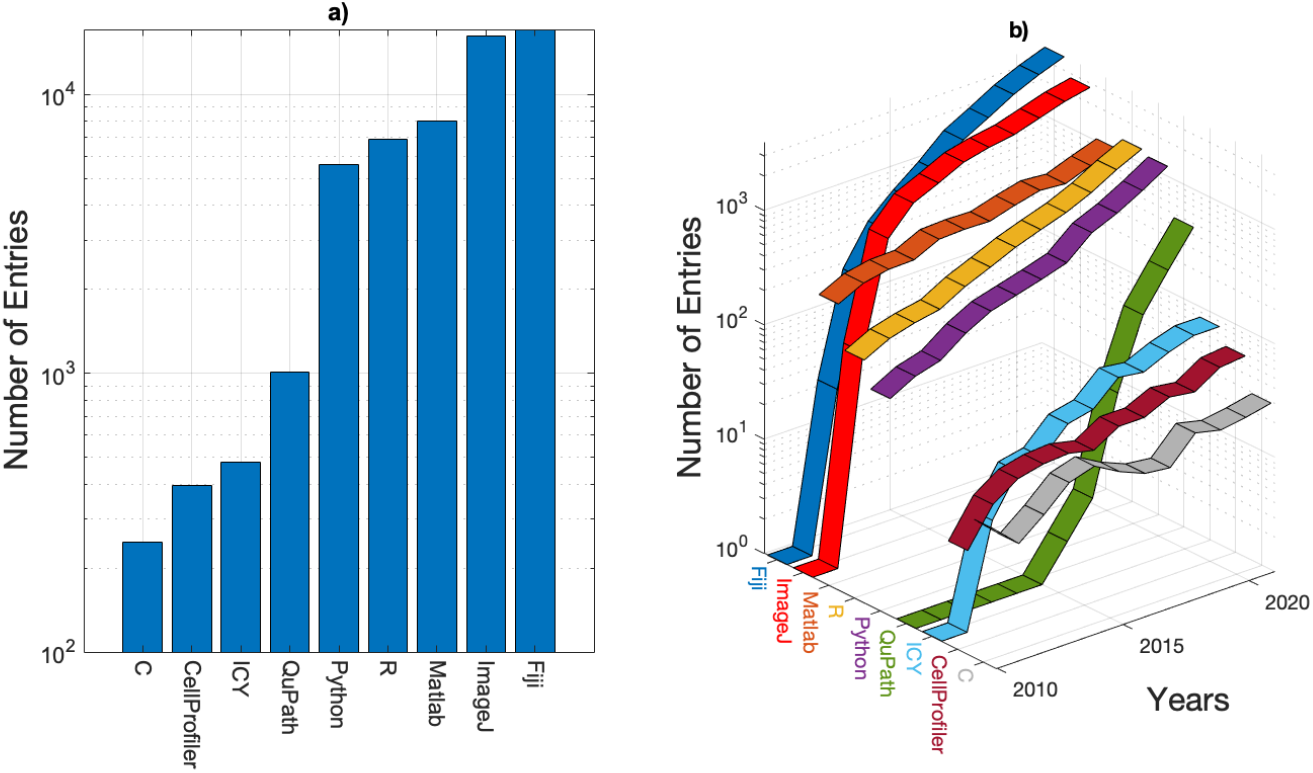
Number of entries in PubMed related to eight programming languages: Fiji, ImageJ, Matlab, R, Python, QuPath, ICY, CellProfiler and C. Subplot a are the entries for the keywords for the programming languages and all the previous keywords *(pathology) AND (cancer) AND ((image) OR (imaging)) AND (2010:2022[dp])* and subplot b is just for the programming languages and the dates. The vertical axis is logarithmic in both graphs.

The lowest number of entries corresponded to *C*. The relatively low numbers could be due to several factors. The most obvious one is that the C programming language is not widely used by those researchers whose work is indexed in PubMed. It may also be that the work created in C may result in larger tools, which go under other names, for instance, ICY is java-based software platform. One important point to notice is that Matlab is the only of these software platforms that requires a license fee. Whilst this fee is significant for industry, academics can obtain it at reduced cost and in many times it is part of the universities site license. However, this could affect the number of users compared with other platforms that do not require license fees.

Yearly entries show how Fiji has been overtaking ImageJ in recent years. Although Fiji and ImageJ are very closely related, and for some people these are considered to be the same (the actual acronym of FIJI is a recursive Fiji Is Just ImageJ), with the addition of a set plugins that facilitate image analysis, in this publication they are being treated separately to identify the actual name being used in the entries that referred to them. A similar trend can be observed with R and Python overtaking Matlab. A very noticeable increase in entries is that related to QuPath, which has grown at a much faster rate than any other language/software. If this trend continues, it can be speculated that it will soon reach the same number of entries as the programming languages and eventually close to to ImageJ and Fiji.

Whilst this review did not consider the actual biological/clinical tasks such as finding and counting nuclei, estimation of cellularity, semantic annotations, anomaly detection or segmenting organs, these could be investigated with the same methodology as described in this work.

## 4 Conclusions

This scoping review provides a panoramic view of the computational techniques that have been applied for the analysis of images produced in Cancer research. The techniques included in the areas of machine learning and deep learning have grown to dominate the recent works indexed in PubMed. Whilst other techniques are still recorded in the records of PubMed, most of these are decreasing their proportions as compared with machine and deep learning.

The tools with which these techniques have been applied, i.e. the software platforms and programming languages showed that general tools through which researchers programme their own solutions are popular, but some specialised softwares like ImageJ and Fiji are widely used, with some newer options like QuPath increasing their position in this area.

## Data Availability

All data produced are available online at GitHub: https://github.com/youssefarafat/Scoping-Review

## Acknowlegements

We acknowledge Dr Robert Noble for the useful discussions regarding this work.

